# Addressing disruptions in childhood routine immunisation services during the COVID-19 pandemic: perspectives and lessons learned from Liberia, Nepal, and Senegal

**DOI:** 10.1101/2021.03.18.21252686

**Authors:** Sameer M Dixit, Moussa Sarr, Daouda M Gueye, Kyle Muther, T. Ruston Yarnko, Robert A Bednarczyk, Adolphus T Clarke, Aliou Diallo, Bonheur Dounebaine, Anna Ellis, Nancy Fullman, Nathaniel Gerthe, Jhalak S Guatam, Kyra Hester, Gloria Ikilezi, Souleymane Mboup, Rajesh Man Rajbhandari, David E Phillips, Matthew C Freeman

**Affiliations:** Center for Molecular Dynamics Nepal (CMDN), Kathmandu, Nepal; Institut de Recherche en Santé de Surveillance Epidémiologique et de Formations (IRESSEF), Dakar, Senegal; Last Mile Health, Monrovia, Liberia; Rollins School of Public Health, Emory University, Atlanta, Georgia, United States; Expanded Programme on Immunisation, Ministry of Health, Monrovia, Liberia; Expanded Programme on Immunisation Unit, World Health Organization Country Office, Dakar, Senegal; Gates Ventures, Kirkland, Washington, United States; Child Health and Immunization Section, Family Welfare Division, Department of Health Services, Ministry of Health and Population, Kathmandu, Nepal

**Keywords:** routine immunisation, COVID-19, childhood vaccines

## Abstract

The COVID-19 pandemic has inflicted multifaceted disruptions to routine immunisation from global to local levels, affecting every aspect of vaccine supply, access, and demand. Since March 2020, country programmes have implemented a range of strategies to either continue vaccination services during COVID-19 measures like ‘lockdown’ and/or resume services when risks of SARS-CoV-2 transmission could be appropriately mitigated. Through the Exemplars in Global Health partnership in Liberia, Nepal, and Senegal, we conducted interviews with immunisation programme managers and ministry of health leadership to better understand how they have addressed the myriad vaccination challenges posed by the ongoing pandemic. From establishing alternative modes of service delivery to combatting vaccine distrust and rumours via risk communication campaigns, many routine immunisation programmes have demonstrated how to adapt, resume, and/or maintain vital vaccination efforts during the COVID-19 crisis. Yet millions of children remain un- or under-vaccinated worldwide, and the same programmes striving to implement catch-up services for missed doses and postponed mass campaigns will also soon be tasked with COVID-19 vaccine deployment. As laid bare by the current pandemic, the world’s gains against vaccine-preventable diseases are fragile: enshrined by a delicate global ecosystem of logistics, supply, and procurement, the success of routine immunisation ultimately rests upon dedicated programme staff, the resources and support available to them, and then the trust in and demand for vaccines by their recipients. Our collective lessons learned during COVID-19 offer insights in programme adaptation and resilience that, if prioritised, could strengthen equitable, sustainable vaccine delivery for all populations.

**Summary box:**

- <u>Key message 1</u>: As the COVID-19 pandemic affected routine immunisation services worldwide, country programmes have used a range of mitigation strategies to maintain vaccine delivery and/or resume interrupted programming. Interviews with immunisation programme managers and Ministry of Health staff provided key perspectives and lessons learned on how countries have approached routine immunisation services during the COVID-19 crisis.
- <u>Key message 2</u>: Key themes for mitigating COVID-19’s effects on routine immunisation included prioritising continued services with strengthened infection prevention control; identifying alternative locations and approaches to providing vaccine services (e.g., conducting door-to-door vaccination if facility-based services were not possible); engaging in effective communications and mobilisation activities, especially to offset misinformation about COVID-19 and vaccines; setting up systems and strategies for reaching children who missed doses amid periods of disruption; and conducting catch-up campaigns as soon as SARS-CoV-2 transmission risks could be minimised.
- <u>Key message 3</u>: The ways in which COVID-19 has affected routine immunisation services have varied over time and across settings, underscoring the importance of contextually-tailored mitigation efforts and adaptation given evolving challenges amid an ongoing pandemic. As countries prepare and initiate roll-out COVID-19 vaccines, it will be vital to avoid one-size-fits-all implementation strategies and support the continuance of routine immunisation services through this next phase of COVID-19 response.

## Introduction

By the time WHO declared SARS-CoV-2 a pandemic on 11 March, 2020,[1] the virus responsible for COVID-19 had claimed more than 4,200 lives and incurred over 118,300 confirmed infections in 114 countries.[2] As of 10 January, 2021, the novel coronavirus has caused over 1.9 million deaths and 88.4 million infections globally,[3] with many countries still actively combatting surging caseloads and deaths. Beyond its direct toll, however, the COVID-19 pandemic has posed both immediate threats and potential longstanding consequences as countries sought to curb disease transmission. These include interrupted education and learning as schools closed for in-person instruction,[4] economic devastation as many business sectors followed suit,[5] and as increasingly recognised globally and nationally,[6] widespread disruptions in vital health services. In August 2020, 90% of 105 countries surveyed by WHO reported at least some kind of disruption to health services during the pandemic.[6] Both demand and supply were disrupted (e.g., outpatient attendance fell amid lockdown measures, many clinics paused operations amid staff redeployment for COVID-19 response). Declines in health service utilisation may have been further fueled by fear of virus exposure and misinformation.[7,8]

Globally, routine immunisation emerged as one of more acutely affected health services,[6] a trend that could result in millions of children un- or under-vaccinated against preventable illness and death if left unaddressed.[9] The very nature of immunisation services – individuals receiving doses from trained vaccinators in close proximity to each other – makes them less directly amenable to telemedicine or other remote delivery options (e.g., medications sent by mail). Further, how countries may approach mitigating pandemic-related disruptions to immunisation services is likely to vary. Factors such as programme structure (e.g., regular use of mass vaccination campaigns and community outreach to complement facility-based services), composition of national immunisation schedules and plans for new vaccine introductions, supply chain requirements and dependencies, and vaccine sentiment may shape strategies to maintain or resume immunisation services amid the ongoing COVID-19 crisis. Together, they present formidable challenges to any country. At the same time, these challenges have also paved the ways for innovation, adaptions, and resilience in the ways that health workers, programme managers, and government leadership alike approach routine immunisation.

To better understand how COVID-19 has affected routine immunisation and the ways in which countries are responding, we used a two-pronged approach. First, we conducted interviews with expanded programme on immunisation (EPI) managers, government officials, and other key stakeholders in Liberia, Nepal, and Senegal. These three countries are part of the Exemplars in Global Health (EGH) programme,[10] an initiative focused on evaluating successes in global health programme implementation and investments, and disseminating lessons learned via partnership networks. The timing and scope of these interviews varied (Table 1), and they were selected by country EGH leadership. As such, they should be interpreted as individual insights into how each country’s routine immunisation programmes have experienced and/or addressed the unique challenges posed by COVID-19 at the time of interview. These perspectives and experiences may not be shared by others across vaccine programmes in a given setting, and they may not represent challenges and/or mitigation strategies currently occurring in these contexts. Second, we synthesised these interviews into key themes across the three countries, supplementing thematic considerations *via* a pragmatic review of publicly available reports and documentation on how COVID-19 was affecting routine immunisation and corresponding mitigation efforts taken by the world more broadly. We primarily drew from individual country reports or data collated by global vaccine partners (e.g., WHO, UNICEF, Gavi, the Vaccine Alliance, Sabin Vaccine Institute) from July to September 2020, with some additional searches in October and November 2020.

**Table 1.** Summary of interviews conducted among routine immunisation progamme and Ministry of Health leadership in Liberia, Nepal, and Senegal. Interviews in Senegal and Nepal were conducted in French and Nepalese, respectively, and were translated into English.

| Country | Interview timing | Interviewee(s) |
| --- | --- | --- |
| Liberia | July-August 2020 | **One interviewee:**<br>• National-level EPI manager, Ministry of Health |
| Nepal | July-August 2020 | **Two interviewees:**<br>• Chief of Child Health and Immunisation Chief at the Family Welfare Division (FWD), Ministry of Health and Population (MoPH)<br>• One district Health Section Chief in charge of immunisation |
| Senegal | September-November 2020 | **Four interviewees:**<br>• Two district-level EPI supervisors<br>• EPI focal point/WHO liaison with the Ministry of Health<br>• District-level physician, national COVID-19 response team |

In the sections that follow we share key insights and themes from Liberia, Nepal, and Senegal, as well as additional learnings to date. It is worth noting that the broader impacts of COVID-19 on routine immunisation may not be wholly known until months or even years from the present crisis. Similarly, the longer-term success of current efforts to maintain or resume critical vaccination services remains unclear; this is particularly true as epidemic trajectories have varied widely across settings over time and now many countries are preparing for COVID-19 vaccine deployment. Here we aim to bring together what has been reported to date; as more data are collected, analysed, and disseminated, we hope to collectively chart the different types of programmatic strategies and innovations that countries implemented to sustain vaccine delivery during the COVID-19 crisis.

### COVID-19 effects on childhood immunisation and country approaches

The COVID-19 pandemic has disrupted key drivers of vaccine utilisation – community access, facility readiness, and intent to vaccinate[11] – as well as how these drivers interact to collectively support timely, effective vaccine services worldwide. On their own, each of these disruptions could negatively affect immunisation services and utilisation – together, they are poised to halt or even reverse the immense progress achieved in vaccine delivery. For instance, the September 2020 Goalkeepers report showed that 2020 coverage estimates of three doses of the diphtheria-tetanus-pertussis vaccine (DTP3) fell to levels last seen in the 1990s – “in other words, we’ve been set back by about 25 years in about 25 weeks.”[5]

Table 2 provides some examples of how these drivers have been affected in Liberia, Nepal, and Senegal, as well as other countries and globally, during the ongoing COVID-19 pandemic. These represent only a portion of the overarching theme surfacing in 2020: disruption of routine vaccine delivery has been widespread, and rapidly developing strategies to provide missed doses and close widening gaps in coverage is crucial.

**Table 2.** Main types of disruptions on childhood vaccination during the COVID-19 pandemic. All country examples and insights were synthesised from interviews conducted by Exemplars in Global Health (EGH) country researchers or via desk research and pragmatic reviews. Vaccine drivers are adapted from the Phillips and colleagues’ framework on determinants of vaccination.[11,12]

| Vaccination driver | Examples of COVID-19 disruptions | Country examples and insights |  |
| --- | --- | --- | --- |
|  |  | From EGH interviews in Liberia, Nepal, Senegal | From desk research and pragmatic reviews |
| **Facility readiness:**<br>health system supply and capacity, via health facilities, to adequately meet demand for vaccine services. | • Delayed shipments of vaccine stocks and key supplies due to travel restrictions and reduced flight availability.[13]<br>• Deployment of facility staff for COVID-19 response and/or emergency management.<br>• Reduced or halted service availability and/or hours of operation.<br>• Inadequate PPE, sanitation and infection control supplies.<br>• Staff shortages due to COVID-19 infections, fear of exposure, and/or burnout. | • Large number of immunisation centers ceased operations and outpatient departments were shut down due to COVID-19 cases among healthcare workers (*Nepal*)<br>• Health facility-based outreach activities were suspended in response to growing vaccine hesitancy, which was fueled by rumours circulating on COVID-19 vaccines being tested on citizens (*Liberia*)<br>• Health centre staff were tasked with COVID-19 management and response, reducing their availability for other services (*Senegal*) | • UNICEF reported a 70-80% reduction in planned vaccine shipments between the weeks of 22 March and 1 May 2020.[13]<br>• Healthcare workers reported delays and/or inability to report to posts due to re-assignments to help with COVID-19 cases (*Philippines*)[14]<br>• Delays in cold-chain equipment optimisation equipment and implementation (*multiple Gavi-eligible countries*)[15] |
| **Intent to vaccinate:**<br>demand for vaccines and/or vaccination services by caretakers that, in the absence of all other barriers, would result in children being vaccinated. | • Fear of virus exposure at healthcare settings. [6]<br>• Instructed to postpone or delay non-urgent healthcare services due to concerns about overburdening healthcare facilities.<br>• Misinformation or rumours on COVID-19 and vaccination safety. | • Substantially reduced patient attendance for vaccine services, and even some refusal for care even when home visits were offered (*Senegal*)<br>• Uptake of routine immunisation services declined amid early days of the country's State of Emergency and curfews (*Liberia*)<br>• Rumours of 'eventual evil' COVID-19 vaccine trial in country (*Senegal*) | • More than 50% of parents reluctant to bring children for immunisation services due to mistrust and rumors (*Côte d'Ivoire*)<br>• Parents delayed children's vaccination schedules due to concerns about COVID-19 and potential transmission in crowded places (*Vietnam*)[16] |
| **Community access:**<br>ability to carry out vaccination via barriers and facilitators between facility readiness and vaccination intent. | • Postponement or cancellation of mass vaccination campaigns, as well as new vaccine introductions.<br>• Restrictions on movement outside of residences and physical access to vaccine services due to social distancing policies and lockdown measures enacted to curb viral transmission.[9] | • Halted measles-rubella campaign in mid-April (*Nepal*)<br>• School closures halted school-based HPV service delivery (*Senegal*)<br>• Community-based outreach activities and mobilisation were prohibited (*Senegal*) | • Between March and April 2020, 27 countries postponed measles immunisation campaigns and 38 postponed mass polio vaccination efforts.[9]<br>• 84% of health facilities reported immunisation services were severely disrupted, with reasons including transport disruptions and school closures due to COVID-19 restrictions (*Indonesia*) [17] |

WHO has provided vital guidance on how to provide immunisation services amid COVID-19 since March 2020,[18–21] of which countries have sought to incorporate and adapt in parallel.[22,23] Although WHO never wavered in its support of routine immunisations, its recommendations have evolved alongside the world’s evidence base on COVID-19 and how to weigh the relative risks of SARS-CoV-2 transmission against the costs of service disruption. In late March, for example, for countries with surging COVID-19 epidemics and without active measles or polio outbreaks, WHO recommendations viewed exposing healthcare workers and community members to this novel coronavirus as greater than temporarily postponing mass immunisation campaigns.[18] As the world’s understanding of COVID-19 improved and the magnitude of health service disruptions began to emerge (e.g., an estimated 80 million children under the age of 1 were thought to be affected interrupted immunisation services by May 2020[9]), WHO issued follow-up guidance on implementing mass vaccination campaigns[19] and principles for maintaining or resuming immunisation activities.[20,21] How countries have then acted – adapting such guidelines for local needs or developing their own mitigation strategies against COVID-19 disruptions based on prior experience or some combination of each – has become an area of great interest for programme managers, implementation scientists, and decision-makers alike.

Based on the interviews conducted in Liberia, Nepal, and Senegal, we report on more detail for each country’s unique challenges – and strategies – for routine immunisation during the COVID-19 pandemic.

### A focus on Liberia: identifying key vaccination barriers, implementing risk communication strategies, and augmenting safety measures and facility capacity for services

As Liberia declared a 3-week state of emergency in April 2020 in response to its COVID-19 epidemic, another health crisis was growing: misinformation on COVID-19 and vaccines. Rumors that “a COVID-19 vaccine would be tested on people seeking routine immunisations” were spreading at a virus-like speed, and were seeding heightened vaccine hesitancy, reported the EPI manager. In parallel, vaccine coverage was falling: nationally, coverage of DTP3 and one dose of measles-containing vaccines (MCV1) was 17% and 20% lower, respectively, from January to April 2020 compared with the same time period in 2019.[24] Outbreaks of vaccine-preventable diseases were occurring in Lofa, Bong, and Sinoe counties, among others.

Concerned that the speed and magnitude of vaccine misinformation could have dire longer-term consequences on its broader immunisation programme, Liberia temporarily halted facility-based outreach services in communities and quickly pivoted toward generating evidence-informed approaches to address these emergent challenges. First, Liberia’s EPI team conducted a vaccine perception study Montessrado and Margibi counties, providing vital information on current obstacles for vaccine uptake. This study identified myths about COVID-19, limited trust in both new and routine vaccines, and delays in seeking vaccination services relative to immunisation schedules as among the countries’ primary barriers. Then, in collaboration with Gavi and other key partners, Liberia’s EPI developed a multi-stage communications plan, mapping key messages to target audiences, communication channels, and impact metrics for short-, medium-, and long-term objectives. These activities are currently being implemented across the country.

Resuming routine immunisation services in Liberia has involved careful protocols, supply acquisition, and catch-up immunisation activities. Drawing from WHO guidance, Liberia’s EPI produced a country-specific Infection Prevention and Control (IPC) Guide and implemented training at facilities for vaccinators to support safe administration practices and reduce exposure risk.[18,25] As of August 2020, eleven counties have received “periodic intensification of routine immunisation,” which involves campaign-oriented vaccination sessions with the purpose of catching-up on missed doses. Importantly, Liberia’s EPI team has been actively working with partners to ensure that vaccinators have access to at least basic personal protective equipment (PPE), which is also vital to strengthening trust among individuals as they return to facilities for services and/or are met by vaccinators *via* community outreach campaigns. EPI has also installed Solar Direct Drive cold chain equipment at facilities, investments that benefit both the resumption of routine immunisation services and potentially preparations for COVID-19 vaccine rollout in 2021. A full rebound in service delivery may take months or longer, but Liberia’s EPI is dedicated to delivering for the country’s children; as reported by the EPI manager, “protection of a child against vaccine preventable diseases is a basic right of a child and as such frantic efforts must be [made] to restore and sustain routine immunisation.”

### A focus on Nepal: maintaining routine immunisation services where possible, resuming vaccination campaigns with heightened safety protocols, and ensuring safe practices amid strong demand for vaccination

Nepal enacted lockdown measures at the end of March 2020 and maintained them through much of July 2020, [26] actions considered necessary to contain the novel coronavirus but not ones without consequences for health services and specifically vaccination. For example, about 50% of the country’s immunisation service centers ceased operations during lockdown, falling from approximately 16,000 nationwide to 8,000. Hospitals, including their outpatient departments, had to shut down frequently from COVID-19 cases among healthcare professionals. A measles-rubella campaign was halted mid-way through April 2020 due to safety concerns, and measles outbreaks in 4 of Nepal’s 77 districts were thought to have stemmed from early disruptions to immunisation services.

Nonetheless, Nepal’s Ministry of Health and Population sought to maintain routine immunisation activities wherever possible, directing its Family Welfare Division (FWD) – the agency responsible for vaccination services – to implement safety protocols and adapt service provision as needed. For instance, the FWD conducted outbreak response immunisation (ORI) in districts affected by measles outbreaks, and strengthened monitoring of both measles and other seasonal vaccine-preventable diseases (e.g., tetanus and encephalitis). Local immunisation committees set up 94 ORI centers in affected districts, aiming to ensure that these services were within a 30-minute walk for the majority of populations. Such rapid set-up and implementation was no small feat, given Nepal’s often remote communities and treacherous terrain amid them; in fact, Nepal’s community health workers are known to walk 2-3 hours between settlements and cover 200 households at a time during vaccine campaigns.[27,28]

The nationwide measles-rubella campaign resumed in June 2020, and Nepal introduced the rotavirus vaccine in early July – all during the extended lockdown period. Introducing the rotavirus vaccine required careful, as well as creative, coordination with Gavi to procure and deliver the vaccines and other supplies *via* aircraft, and then to support maximum safety and infection control at immunisation sessions known for their “high-crowd pulling power.”[29] Community vaccine demand and acceptance is very high in Nepal, an achievement associated with the country’s longstanding community health worker programmes (e.g., Female Community Health Volunteers) and strong cultural norms around vaccination. “Despite lockdown, large numbers of [the] public showed up at immunisation centers that were open to vaccinate their children,” reported the Child Health and Immunisation Chief at the FWD. To safely continue routine immunisation amid such high demand, the Nepal government developed PPE guidelines, and ensured that all vaccinators received masks and eyewear, and full PPE whenever possible (i.e., gloves, footwear, overalls in addition to masks and eyewear). Formal training on IPC was conducted and distancing measures were implemented prior to field mobilisation efforts as campaigns resumed.

### A focus on Senegal: overcoming disruptions in vaccination demand and community mobilisation with effective communications and training, locally-adapted guidelines and safety practices, catch-up outreach services

In spring 2020, Senegal’s health workforce was busy not only responding to COVID-19 demands but also adapting WHO guidelines to local needs and implementing sanitation and safety protocols to ensure continued essential service delivery at health centers and clinics. Facility staff developed IPC protocols, established physical distancing measures and procured PPE, and ran or participated in training webinars coordinated with national-level programmes to prepare for immunisation service delivery during the pandemic. During these preparations and protocol implementation, however, key challenges surfaced: in addition to facility staff being deployed COVID-19 management, demand for vaccine services plummeted and Senegal’s usual delivery platforms and mobilisation activities for routine immunisations were no longer viable amid lockdown measures.

Schools were closed, halting HPV vaccine delivery as schools are Senegal’s main platform for HPV vaccination. Community-based organisations (CBOs) and multiple types of community health workers serve as the backbone to many of Senegal’s primary care and child health services, and have been recognised as vital to Senegal’s child health successes. But during the COVID-19 crisis, community mobilisation activities – such as that of the *Badiènou Gokh* to promote vaccination via music and in-person communication campaigns – and outreach-based services were prohibited. Patient attendance at facilities also substantially declined, likely from a combination of restrictions of movement and concerns about COVID-19 exposure. “Patients had deserted the health centers for fear of being infected….we could stay for a week without receiving a patient,” said one interviewee, an immunisation supervisor. Staff would call families and arrange for community workers to visit their homes with full protective equipment, yet in some cases, rumors about COVID-19 and vaccine reliability prompted families to still refuse immunisation services.

To overcome these challenges, Senegal’s routine immunisation staff are re-starting mobilisation efforts with careful compliance to IPC, reduced numbers of individuals at any given time, and proper physical distancing measures. Such work has been informed by the Ministry of Health and Social Actions (MHSA) instructions, which was sent to all regions and health centers, and tailoring existing guidelines to align with local needs and strategies across the country’s health system levels. Media campaigns and broadcasting from cable channels, local TV channels, and radio spots are emphasising the importance of childhood immunisation as EPI programmes are once more scaling up outreach services. Efforts to implement continuous vaccination services outside of health posts in collaboration with CBOs are seeing some success, especially as clinics and outreach programmes have sought to provide greater flexibility in the timing of visit scheduling and location for catch-up doses. In addition, district-level records are enabling clinic staff to send text reminders or phone caretakers of children who have missed scheduled immunisation visits. As of November 2020, interviewees indicated that school-based HPV vaccination has yet to resume; however, they also report that community health workers have begun house-to-house vaccination efforts against HPV. While many obstacles persist, especially as the world continues to battle COVID-19, the dedication of Senegal’s health workers to identifying and implementing solutions for reaching every child was a unifying thread across these interviews. “Let’s immunise our children, the vaccines are there….The government has done everything to make these vaccines available for us, so let’s immunise our children. This is my only cry from the heart” one interviewee, a health facility EPI supervisor, said as their final statement.

### Key mitigation themes and implications for COVID-19 vaccine rollout

As highlighted by Liberia, Nepal, and Senegal, the COVID-19 pandemic has affected routine immunisation services differently across countries – and they, in turn, have approached maintaining and/or resuming vaccination services in myriad ways as well. None of these findings should be surprising, especially to immunisation programme managers, ministry leadership, and vaccine partners – the complexities of vaccine delivery and how they intersect with deeply held sociocultural mores, political commitment, and technical requirements make locally-relevant and tailored programmes fundamental to any successful immunisation effort. Yet at a time when the world is rapidly pivoting to COVID-19 vaccine roll-out, we – collectively – risk viewing such efforts in a one-size-fits-all mentality and losing vital lessons learned on vaccine service provision during the earlier phases of the COVID-19 pandemic.

Specific logistics and implementation approaches for COVID-19 vaccine roll-out will likely deviate from routine immunisation services, ranging from the populations targeted and phases of prioritisation to cold-chain and dosing schedules. At the same time, COVID-19 vaccines will be scaled up amid ongoing, if not rising, SARS-CoV-2 transmission in many countries, and similar to routine immunisation, multifaceted mitigation, communications, and demand-generation activities will be necessary to ensure optimal service provision and safety for all. Table 3 aims to summarise some of the key mitigation themes across Liberia, Nepal, and Senegal, in addition to supplementary examples from other locations. Not every approach was used or needed in each country, just as COVID-19 affected routine immunisation in different ways across settings. What unites these diverse settings and programmes is the recognition that each level of vaccination barriers or facilitators must be addressed, from patient demand and trust to community access; the willingness to adapt and test alternative approaches; and the overarching dedication to providing routine immunisation to all.

**Table 3.** Main mitigation themes for addressing COVID-19 effects on routine immunisations. All country examples and insights were synthesised from interviews conducted by Exemplars in Global Health (EGH) country researchers or via desk research and pragmatic reviews.

|  | Country examples and insights |  |
| --- | --- | --- |
|  | From EGH interviews in Liberia, Nepal, Senegal | From desk research and pragmatic reviews |
| **Theme 1:** Prioritise continued immunisation services at both national and local levels. | Supported uninterrupted routine immunisation services by national government and local authorities, even during lockdown periods (*Nepal, Senegal*)<br>Introduced rotavirus vaccine in July 2020, during a national lockdown period (*Nepal*) | Conducted routine immunisations at fixed sites with physical distancing during national lockdown (*Laos*)[9]<br>Sought to maintain routine immunisation services via strong engagement with religious and community leaders and trainings on vaccine-preventable disease surveillance for volunteers (*Afghanistan*)[30] |
| **Theme 2:** Conduct effective communications and outreach efforts to combat misinformation and engender trust in COVID-19 guidance and routine immunisation efforts. | Developed and now currently implementing risk communication strategic action plan to address widespread rumors and vaccine hesitancy identified through a vaccine perception study (*Liberia*)<br>Conducted communication campaigns and mobilisation efforts via multiple media channels emphasising the importance of childhood vaccines (*Senegal*) | Conducted virtual campaigns with the President and Minister of Health via social media (*Paraguay*)[31]<br>Dispatched mobile teams of community health workers with longstanding ties to communities targeted for outreach (*Cambodia*) [32]<br>Ministry of Health leveraged mass media to inform populations that essential health services had resumed, including vaccination, and urged community members to bring children to clinics (*Sri Lanka*) [33] |
| **Theme 3:** Identify alternative locations, infrastructure, and service delivery platforms outside of traditional health clinics. | Offered alternative hours for vaccine administration (e.g., after work, during the weekends) (*Senegal*)<br>Set up outbreak response immunisation centers in districts with outbreaks to facilitate faster catch-up and response immunisation (*Nepal*) | Set up vaccination posts in empty locations due to lockdown measures (e.g., schools) (*Brazil*)[31]<br>Conducted drive-through and home-based vaccination services (*Brazil, Chile, El Salvador*)[31]<br>Deployed “immunisation brigades” to high-risk populations at nursing homes and jails (*Bolivia*)[31] |
| **Theme 4:** Institute infection prevention controls and physical distancing measures, and adapt service provision practices accordingly. | Developed locally tailored IPD guidelines and trainings for vaccinators (*Liberia, Senegal*)<br>Improved the provision of basic PPE and other supplies for vaccinators via country partners (*Liberia*) | Trained polio vaccine workers to administer vaccines without touching children, alongside provision of full PPE (*Pakistan*)[34]<br>Introduced rotavirus vaccine via a phased province-by-province approach rather than the centralised introduction model, advising people not to travel outside of their home provinces to reduce transmission risk (*Solomon Islands*)[35]<br>Repurposed government cars into shuttles to drive patients and caregivers to clinics to access health services including immunisations (*Uganda*)[30]<br>Limited patients to one child or family per hour for immunisation services, and sought to give multiple catch-up vaccinations and/or vaccinate all children in a given family per visit when possible (*Sri Lanka*) [32,33]<br>Set up tents outside of health centers to separate people seeking vaccination from patients who are ill (*Paraguay*)[31] |
|  |  | Extended mass measles campaign of 14.4 million children to a longer time period to limit crowding, and conducted vaccinations in outside, well-ventilated areas with physical distancing and sanitation protocols (*Ethiopia*)[36] |
| **Theme 5:** Set up systems to track missed doses – and target follow-up efforts – in areas with the largest disruptions. | Conduct house-to-house vaccination for HPV services amid continued closures of schools (*Senegal*)<br>Based on contact records kept at district offices, text or phone parents whose children have missed immunisation visits (*Senegal*) | Tracked children who missed doses and sent text messages about service resumption via its Electronic Immunisation Registry (EIR) mobile application (*Karachi, Pakistan*)[37]<br>Phoned parents of children with missed vaccination and scheduling home visits as needed (*Ghana*)[38] |
| **Theme 6:** Conduct catch-up campaigns as soon as SARS-CoV-transmission risks can be effectively mitigated. | Conducted “periodic intensification of routine immunisation” (i.e., campaign-style vaccine services) in at least eleven districts by August 2020 (*Liberia*)<br>Resumed measles-rubella mass vaccination campaign in June 2020 after halting activities in mid-April due to safety concerns (*Nepal*) [32] | Resumed postponed polio vaccination campaigns with extensive PPE and safety protocols, including Syria in June,[28,32] Burkina Faso,[39] and Pakistan[34] in July, among others.<br>Leveraged June 2020 Child Health Week to include catch-up campaigns for inactivated polio vaccines and HPV vaccination (*Zambia*)[35] |

## Conclusions

The COVID-19 pandemic has threatened progress made in child health worldwide, posing widespread disruptions and challenges to routine immunisation programmes, healthcare providers, and patients alike. Programmatic lessons learned through this global crisis are only as beneficial as our collective efforts to collaborate and rethink approaches to promoting programme resilience, equity, and sustainability for both immunisation and preparedness more broadly. In the coming months, every country – rich and poor, Global South and North – will face its next reckoning of COVID-19: mass deployment of COVID-19 vaccines amid ongoing, or even surging, epidemics and SARS-CoV-2 transmission. How we collectively approach routine immunisation services and vaccine campaigns, from local to global levels, will ultimately determine how the remainder of this pandemic will unfold.

## Data Availability

Requests for de-identified interview data will be reviewed by country team leads; all other data referred to in the manuscript are publicly available.

## Acknowledgments

We are deeply grateful for the time and responses provided by all interviewees and interviewers, especially as they conduct vital work amid the COVID-19 pandemic. We would like to acknowledge Nepal’s Ministry of Health and Population and the WHO Nepal Immunization Preventable Diseases section; Senegal’s Ministry of Health and Social Actions and the WHO Country Office in Senegal; and Liberia’s Ministry of Health. The Exemplars in Global Health programme is a partnership including the institutional affiliations of the authors listed here as well as country research partners. The authors acknowledge contributions of all partners involved, which also include country leadership overseeing research collaborations in Liberia, Nepal, and Senegal; Anna Rapp and Ethan Wong of the Bill & Melinda Gates Foundation; and Daniel Beaulieu, Niranjan Bose, and Oliver Rothschild at Gates Ventures. Additionally, we thank Angela Hwang for her critical perspectives and feedback on routine immunisation programmes and drafting of the manuscript. The Exemplars in Global Health Vaccine Delivery project, coordinated by Emory University, is funded by the Bill & Melinda Gates Foundation in conjunction with Gates Ventures.

## References

1 Cucinotta D, Vanelli M. WHO Declares COVID-19 a Pandemic. Acta Bio-Medica Atenei Parm 2020;91:157–60. doi:10.23750/abm.v91i1.9397

2 World Health Organization (WHO). Coronavirus disease 2019 (COVID-19): Situation Report 51. Geneva, Switzerland: : WHO 2020. https://www.who.int/docs/default-source/coronaviruse/situation-reports/20200311-sitrep-51-covid-19.pdf?sfvrsn=1ba62e57_10

3 World Health Organization (WHO). COVID-19 Weekly Epidemiological Update: 12 January 2021. Geneva, Switzerland: : WHO 2021. https://www.who.int/publications/m/item/weekly-epidemiological-update---12-january-2021 (accessed 13 Jan 2021).

4 UNICEF. What Have We Learnt? Overview of findings from a survey of ministries of education on national responses to COVID-19. New York, NY, USA: : UNICEF 2020. https://data.unicef.org/resources/national-education-responses-to-covid19/

5 Bill & Melinda Gates Foundation (BMGF). 2020 Goalkeepers Report. COVID-19: A Global Perspective. Seattle, WA, USA: : BMGF 2020. https://www.gatesfoundation.org/goalkeepers/report/2020-report

6 World Health Organization (WHO). Pulse survey on continuity of essential health services during the COVID-19 pandemic: interim report. Geneva, Switzerland: : WHO 2020. https://www.who.int/publications-detail-redirect/WHO-2019-nCoV-EHS_continuity-survey-2020.1 (accessed 4 Sep 2020).

7 World Health Organization (WHO). Novel Coronvirus (2019-nCoV): Situation Report 13. Geneva, Switzerland: : WHO 2020. https://www.who.int/docs/default-source/coronaviruse/situation-reports/20200202-sitrep-13-ncov-v3.pdf?sfvrsn=195f4010_6

8 The Risks of Misinformation and Vaccine Hesitancy within the Covid-19 Crisis. https://www.csis.org/analysis/risks-misinformation-and-vaccine-hesitancy-within-covid-19-crisis (accessed 4 Sep 2020).

9 UNICEF. At least 80 million children under one at risk of diseases such as diphtheria, measles and polio as COVID-19 disrupts routine vaccination efforts, warn Gavi, WHO and UNICEF. 2020.https://www.unicef.org/press-releases/least-80-million-children-under-one-risk-diseases-such-diphtheria-measles-and-polio (accessed 3 Aug 2020).

10 Exemplars in Global Health. exemplars.health. https://www.exemplars.health/ (accessed 3 Sep 2020).

11 Phillips DE, Dieleman JL, Lim SS, et al. Determinants of effective vaccine coverage in low and middle-income countries: a systematic review and interpretive synthesis. BMC Health Serv Res 2017;17:681. doi:10.1186/s12913-017-2626-0

12 Phillips DE, Dieleman JL, Shearer JC, et al. Childhood vaccines in Uganda and Zambia: Determinants and barriers to vaccine coverage. Vaccine 2018;36:4236–44. doi:10.1016/j.vaccine.2018.05.116

13 UNICEF. Geneva Palais briefing note on the impact of COVID-19 mitigation measures on vaccine supply and logistics. 2020.https://www.unicef.org/press-releases/geneva-palais-briefing-note-impact-covid-19-mitigation-measures-vaccine-supply-and (accessed 3 Aug 2020).

14 BrandRoom I net. Missed a vaccine? It’s time to bounce back! INQUIRER.net. 2020.https://globalnation.inquirer.net/190451/missed-a-vaccine-its-time-to-bounce-back (accessed 11 Dec 2020).

15 Gavi, the Vaccine Alliance. COVID-19 Situation Report 8. Geneva, Switzerland: : Gavi 2020. https://www.gavi.org/sites/default/files/document/2020/Gavi-COVID-19-Situation-Report-8-05052020.pdf

16 COVID-19 Situation Report 10. Geneva, Switzerland: : Gavi 2020. https://www.gavi.org/sites/default/files/document/2020/Gavi-COVID-19-Situation-Report-10-20200602.pdf

17 UNICEF. Rapid Assessment: Impact of COVID-19 Pandemic on Immunization Services in Indonesia. New York, NY, USA: : UNICEF 2020. https://www.unicef.org/indonesia/media/4811/file/Rapid%20Assessment:%20Impact%20of%20COVID-19%20Pandemic%20on%20Immunization%20Services%20in%20Indonesia.pdf

18 World Health Organization (WHO). Guiding principles for immunization activities during the COVID-19 pandemic: interim guidance. Geneva, Switzerland: : WHO 2020. https://www.who.int/publications-detail-redirect/guiding-principles-for-immunization-activities-during-the-covid-19-pandemic-interim-guidance (accessed 3 Aug 2020).

19 World Health Organization (WHO). Framework for decision-making: implement of mass vaccination campaigns in the context of COVID-19. Interim guidance. Geneva, Switzerland: : WHO 2020. https://apps.who.int/iris/bitstream/handle/10665/332159/WHO-2019-nCoV-Framework_Mass_Vaccination-2020.1-eng.pdf

20 World Health Organization (WHO). Maintaining essential health services: operational guidance for the COVID-19 context interim guidance. Geneva, Switzerland: : WHO 2020. https://www.who.int/publications/i/item/WHO-2019-nCoV-essential-health-services-2020.1

21 World Health Organization (WHO). Immunization as an essential health service: guiding principles for immunization activities during the COVID-19 pandemic and other times of severe disruption. Geneva, Switzerland: : WHO 2020. https://www.who.int/publications/i/item/immunization-as-an-essential-health-service-guiding-principles-for-immunization-activities-during-the-covid-19-pandemic-and-other-times-of-severe-disruption

22 TechNet. TechNet Resource Library on immunization resources. TechNet-21. https://www.technet-21.org/en/library?t=222-covid-19 (accessed 4 Aug 2020).

23 PATH. COVID-19 Essential Health Services Policy Tracker Dashboard. https://tableau.path.org/t/BMGF/views/COVID-19EHSPolicyTrackerDashboard/Welcome?:isGuestRedirectFromVizportal=y&:embed=y

24 Last Mile Health. EPI RED Categorization Tool for Liberia [dataset]. 2020.

25 Expanded Programme on Immunization, Ministry of Health. Infection Prevention and Control Guide for Vaccinators. Liberia: : EPI, MOH 2020.

26 Nepali Times. Nepal ends COVID-19 lockdown. https://www.nepalitimes.com/latest/nepal-ends-covid-19-lockdown/ (accessed 11 Dec 2020).

27 Kandel N, Lamichhane J. Female health volunteers of Nepal: the backbone of health care. The Lancet 2019;393:e19–20. doi:10.1016/S0140-6736(19)30207-7

28 Gavi, the Vaccine Alliance. Delivering life-saving vaccines during the COVID-19 pandemic. 2020.https://www.gavi.org/vaccineswork/delivering-life-saving-vaccines-during-covid-19-pandemic (accessed 7 Aug 2020).

29 World Health Organization (WHO). Nepal introduces Rota virus vaccine against diarrhoea in children: National Immunization Programme achieves new milestone. https://www.who.int/nepal/news/detail/02-07-2020-nepal-introduces-rota-virus-vaccine-against-diarrhoea-in-children-national-immunization-programme-achieves-new-milestone (accessed 4 Aug 2020).

30 Gavi, the Vaccine Alliance. COVID-19 Situation Report 9. Geneva, Switzerland: : Gavi 2020. https://www.gavi.org/sites/default/files/document/2020/Gavi-COVID-19-Situation-Report-9-05192020_0.pdf

31 Pan American Health Organization (PAHO). PAHO urges countries to maintain vaccination during COVID-19 pandemic. 2020.http://www.paho.org/en/news/26-4-2020-paho-urges-countries-maintain-vaccination-during-covid-19-pandemic (accessed 4 Aug 2020).

32 World Health Organization (WHO). How WHO is supporting ongoing vaccination efforts during the COVID-19 pandemic. 2020.https://www.who.int/news-room/feature-stories/detail/how-who-is-supporting-ongoing-vaccination-efforts-during-the-covid-19-pandemic (accessed 4 Aug 2020).

33 Putting Women and Children First: Immunization Resumes in Sri Lanka amidst the COVID-19 Pandemic. https://www.who.int/southeastasia/news/feature-stories/detail/putting-women-and-children-first-immunization-resumes-in-sri-lanka-amidst-the-covid-19-pandemic (accessed 11 Dec 2020).

34 Gadzo M. Pakistan resumes polio vaccinations after coronavirus hiatus. https://www.aljazeera.com/news/2020/07/pakistan-resumes-polio-vaccinations-coronavirus-hiatus-200720071328769.html (accessed 4 Aug 2020).

35 Gavi, the Vaccine Alliance. COVID-19 Situation Report 13. Geneva, Switzerland: Gavi 2020. https://www.gavi.org/sites/default/files/covid/Gavi-COVID-19-Situation-Report-13-20200714.pdf

36 Ethiopia vaccinates nearly 15 million children against measles despite COVID-19 challenges. WHO Reg. Off. Afr. https://www.afro.who.int/news/ethiopia-vaccinates-nearly-15-million-children-against-measles-despite-covid-19-challenges (accessed 11 Dec 2020).

37 Chandir S, Siddiqi DA, Setayesh H, et al. Impact of COVID-19 lockdown on routine immunisation in Karachi, Pakistan. Lancet Glob Health 2020;0. doi:10.1016/S2214-109X(20)30290-4

38 Ghana’s community nurses deliver child health care amid COVID-19. WHO Reg. Off. Afr. https://www.afro.who.int/news/ghanas-community-nurses-deliver-child-health-care-amid-covid-19 (accessed 11 Dec 2020).

39 Burkina Faso resumes polio vaccination campaigns under strict COVID-19 prevention measures. WHO Reg. Off. Afr. https://www.afro.who.int/news/burkina-faso-resumes-polio-vaccination-campaigns-under-strict-covid-19-prevention-measures (accessed 11 Dec 2020).

